# Shielding of Viruses such as SARS-CoV-2 from Ultraviolet Radiation in Particles Generated by Sneezing or Coughing: Numerical Simulations of Survival Fractions

**DOI:** 10.1101/2021.03.23.21254172

**Authors:** Steven C. Hill, David C. Doughty, Daniel W. Mackowski

## Abstract

SARS-CoV-2 and other microbes within aerosol particles can be partially shielded from UV radiation. The particles refract and absorb light, and thereby reduce the UV intensity at various locations within the particle. Shielding has been demonstrated in calculations of UV intensities within spherical approximations of SARS-CoV-2 virions that are within spherical particles approximating dried-to-equilibrium respiratory fluids. The purpose of this paper is to calculate the survival fractions of virions (i.e., the fractions of virions that can infect cells) within spherical particles approximating dried respiratory fluids, and to investigate the implications of these calculations for using UV light for disinfection. The particles may be on a surface or in air. In this paper the survival fraction (*S*) of a set of virions illuminated with a UV fluence (*F*, in J/m^2^) is approximated as *S=*exp(*-kF)*, where *k* is the UV inactivation rate constant (m^2^/J). The average survival fractions (*S*_p_) of all the simulated virions in a particle are calculated using the calculated decreases in fluence. The results show that virions in particles of dried respiratory fluids can have significantly larger *S*_p_ than do individual virions. For individual virions, and virions in 1-, 5-, and 9-µm particles illuminated (normal incidence) on a surface with 260-nm UV light, the *S*_p_ = 0.00005, 0.0155, 0.22 and 0.28, respectively, when *kF=*10. The *S*_p_ decrease to <10^−7^, <10^−7^, 0.077 and 0.15, respectively, for *kF*=100. Calculated results also show that illuminating particles with UV beams from widely separated directions can strongly reduce the *S*_p_. These results suggest that the size distributions and optical properties of the dried particles of virion-containing respiratory fluids are likely important in effectively designing and using UV germicidal irradiation systems for microbes in particles. The results suggest the use of reflective surfaces to increase the angles of illumination and decrease the *S*_p_. The results suggest the need for measurements of the *S*_p_ of SARS-CoV-2 in particles having compositions and sizes relevant to the modes of disease transmission.

## INTRODUCTION

Some diseases caused by viruses and bacteria can be transmitted by sneezed or coughed aerosols, droplets (of any size), or by particles remaining after droplets of respiratory or other fluids have dried-to-equilibrium (termed “dried particles” here) (Lu et al., 2020; Miller et al., 2020; Huang et al., 2021). SARS-CoV-2 is found in fluids of the nasopharynx (Landry et al., 2020; Yilmaz et al., 2021), nose (Pere et al., 2020), throat (Zou et al., 2020), and lung (Wolfel et al., 2020), and in saliva (Azzi et al., 2020; Wyllie et al., 2020) and sputum (Pan et al., 2020). These fluids may be aerosolized by coughing, sneezing, talking or breathing. Microbes in particles on surfaces may be transferred to humans by direct contact, via fomites, or may be reaerosolized (Fisher et al., 2012; Kesavan et al., 2017; Krauter et al., 2017; Paton et al., 2015; Qian et al., 2014). An approach to reducing transmission of the associated diseases is to reduce the exposure of persons to viable microbes in the air and/or on surfaces (Cheng et al., 2020). There is a need for improved designs of buildings and vehicles, and the airflows within them, to reduce the transmission of airborne diseases while allowing human interactions to the extent possible (Morawska et al., 2020; Augenbraun et al., 2020). There is a need for decontamination of masks and filters (Fischer et al., 2020; Woo et al., 2012), and surfaces in many locations. UV germicidal irradiation (UVGI) appears to be useful in addressing these needs (Rutala and Weber, 2019; Lindsley et al., 2018; Morawska et al., 2020; Kowalski, 2009). Here, following Kowalski (2009), both 260- and 302-nm light are included in UVGI.

A problem with using UVGI to inactivate microbes is that in certain cases viruses and bacteria can be partially shielded from UVGI by being within or on particles (Osman et al., 2008; Kesavan et al., 2014; Handler and Edmonds, 2015; Doughty et al., 2021) or by clumping of particles (Coohill and Sagripanti, 2008). Numerical simulations of spherical dried droplets of respiratory fluids containing spherical 100-nm virions to simulate SARS-CoV-2 showed that virions in dried particles of respiratory fluids can be partially protected from UV radiation (Doughty et al., 2021). That study showed that partial shielding from UV occurs in particles whether they are on a surface or not. In these model results the electromagnetic field equations (Maxwell’s equations) were solved exactly (within numerical error) using the Multi-Sphere T-Matrix (MSTM) method (Mackowski, 2008; Mackowski and Mishchenko, 2011, 2013).

In such simulations, virions can be protected both by absorption of UVGI within the particle and by refraction at the air-particle surface. In these simulations the extent of shielding is affected by the wavelength of the UV light and the properties of the dried droplet containing the virion. The degree of shielding increases as the size of the particle containing the virions increases. Illumination from multiple incident beams, from widely separated directions, tends to decrease the shielding from UVGI. Illumination of a particle uniformly from all directions can reduce the shielding. Such all-direction illumination may occur for a particle in turbulent air illuminated for a sufficient time to tumble and spin through all orientations relative to the incident beam(s). The key parameter calculated was the *R*_*i*_, which is the ratio of the UV intensity in the *i*^th^ virion in a particle to the intensity in an individual virion, illuminated either in air or on the same surface.

In modeling and designing systems and procedures for minimizing disease transmission, the survival fraction is typically a more useful quantity than *R*_*i*_. The simplest common expression for survival fraction (*S)* is

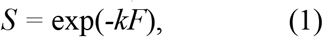

where *k* is the UV rate constant for inactivation (m^2^/J); *F* is the fluence (J/m^2^) (also termed the dose). Also, *F=It*, where *I* is the intensity of the irradiance (W/m^2^), and *t* is the time of illumination. The value of *S* depends on several factors including the UV-intensity at each virion, UV wavelength, time of illumination, humidity, temperature, material in which the virions are embedded (if any), and surface on which the virions rest (if any). For a given virus and set of experimental conditions the *S* is measured for different *F*, and then *k* is determined using Eq. 1. The *k* have been measured for coronaviruses (Hessling et al., 2020), including SARS-CoV-2 (Heilingloh et al., 2020; Ratnesar-Shumate et al., 2020), and other viruses (Sagripanti and Lytle, 2011; Schuit et al., 2020), bacteria and fungi (Kowalski, 2009). More complex expressions of the survival fraction, such as those which incorporate an additional term or terms to account for shoulders, long tails or other features in survival curves (Kowalski 2009, Ch. 3; Kowalski et al., 2019) are beyond the scope of this paper.

Here the average survival fractions are calculated for virions of SARS-CoV-2 (approximated as 100-nm spherical particles) within otherwise homogeneous spherical particles having optical properties to approximate dried particles of sneezed or coughed respiratory fluids. The particles, incident beams, wavelengths and surfaces are as in Doughty et al. (2021): three sizes (1-, 5- and 9-µm), two UV wavelengths (260 and 302 nm), and several combinations of incident beams and directions. Also, the particles are on a surface or in air. The goal of this work is to quantify the increases in survival of virions in model particles that result from the particle’s partial shielding of some of the virions from UVGI. Once users of germicidal- or solar-UVGI are aware of the potential for increases in survival fractions of virions in particles, they may be better able to mitigate the effects of such shielding.

## METHODS

The methods of calculating the UV intensities and estimating the optical properties of the virions and dried respiratory fluids were described in Doughty et al. (2021). If a UV planewave propagates in a material with complex refractive index *m*_*r*_ *+ i m*_*i*_, the intensity of the irradiance of the wave at position (*z*) is *I(z) = I*_*o*_ exp(*-z/δ)*. Here, *I*_*o*_ is *I* at *z=*0, *δ* is the penetration depth, and the imaginary part of the complex refractive index is *m*_*i*_ = *λ/4πδ*, where *λ* is the UV wavelength. The UV intensities in the spherical particles were obtained by solving exactly (within numerical error) the electromagnetic (EM) field equations for a number of homogeneous spheres, either separate or included within other spheres. There may be a planar surface near the sphere(s). No sphere surfaces can intersect. The solution, obtained using the Multi-Sphere T-Matrix (MSTM) method (Mackowski, 2008; Mackowski and Mishchenko, 2011, 2013), expands the fields within each sphere as a sum of vector spherical harmonics (VSH) with unknown coefficients and within the material of the surface using an angular spectrum of planewaves. The coefficients of the VSH and plane waves are obtained by enforcing the boundary conditions at each interface. The virions were approximated as homogenous 100-nm diameter spheres with optical properties chosen to approximate SARS-CoV-2 virions. The enclosing spheres have optical properties selected to approximate dried respiratory fluids as described in Doughty et al., (2021). For illumination with a plane wave having an intensity of irradiance *I*_o_, the net rate of energy absorption by the *i*^th^ virion is 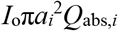, where *a*_*i*_ is the radius and *Q*_abs,*i*_ is the absorption efficiency, each for the *i*^th^ virion. The relative rate of absorption of UV energy of the *i*^th^ virion is *R*_*i*_ = Q_abs,i_ / Q_abs,single_, where Q_abs,single_ is the absorption efficiency of a virion resting on the surface. The *R*_*i*_ can range over several orders of magnitude in absorbing particles with dimensions of several wavelengths. In such cases, large numbers of *R*_*i*_ are needed to adequately simulate the distribution. For 9-µm particles, 120 simulations of 100 virions/particle were calculated and combined to approximate the *R*_*i*_ distribution for randomly positioned virions.

The survival fraction for the *i*^th^ virion in a particle is then

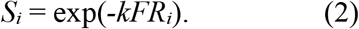

The average of the survival fractions (*S*_*i*_) of all the modeled virions in the particle is *S*_p_, the key parameter illustrated in the figures.

## RESULTS

### Average Survival Fractions are Larger in Larger Particles

Figure 1 shows *S*_p_ (average of the *S*_*i*_ in Eq. 2) as a function of *kF* for virions within 1-, 5- and 9-µm diameter particles, and for individual virions (particle size = virion size = 0.1 µm) as in Eq. 1. In Fig. 1 the *S*_p_ increases with particle size for both wavelengths used (260 and 302 nm). For each of the curves in Fig. 1 (and in Figs. 2 to 4), the Table gives the *S*_p_ at *kF* = 1, 10, 100, 1000 and 10,000. In the figures and the Table, the *S*_p_ is shown as a function of the dimensionless parameter *kF*: for a relevant combination of *k* and *F*, the *S*_p_ can be estimated for particles of the sizes and complex refractive indexes used here. As seen in Fig. 1 and the Table, when λ=260 nm and *kF*=10 the *S*_p_ is: 0.000045 for the 0.1-µm diameter individual virion, 0.015 for the 1-µm particles, 0.217 for 5-µm particles, and 0.28 for the 9-µm particles. When λ=260 nm and *kF*=100 the *S*_p_ is: < 0.0000005 for the 1-µm particles, 0.077 for the 5-µm particles, and 0.149 for the 9-µm particles. The curves in Figs. 1-4 are not simple exponentials (except for the individual-virion curves) because they are averages of many simple exponentials. For the *i*^th^ exponential curve, *S*_*i*_ = *1/e* when *kF* = 1/*R*_*i*_. As *kF* increases, the *S*_*i*_ of the virions with the largest *R*_*i*_ decrease faster than do the *S*_*i*_ of virions with smaller *R*_*i*_. Consequently, the relative contributions of the different virions to the *S*_p_ vs *kF* curve changes with *kF*. For large *kF* the virions with the smallest *R*_*i*_ dominate the *S*_p_.

**Figure 1.**
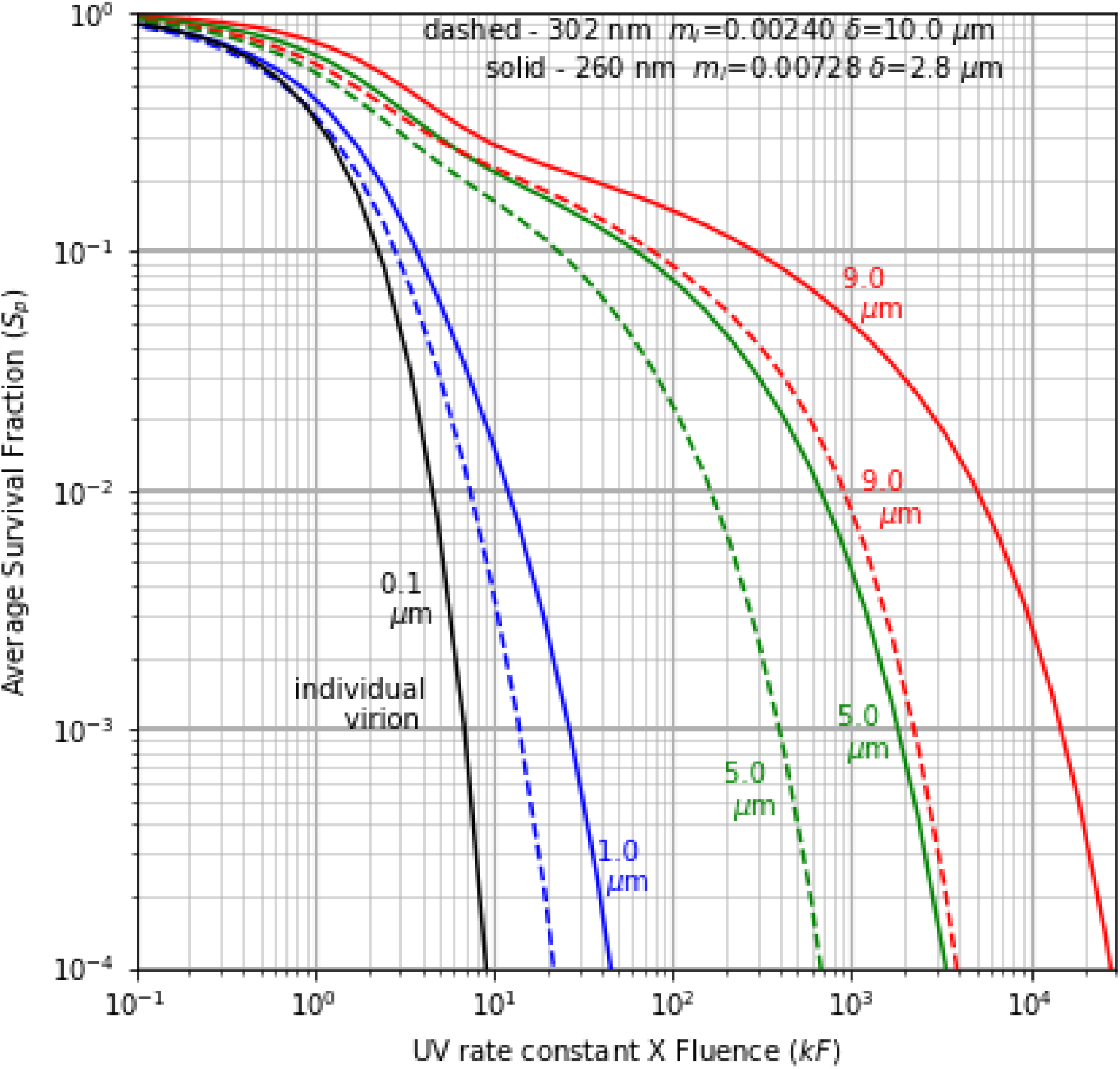
Average survival fractions (*S*_p_) vs *kF*, the product of the UV rate constant (*k*) and the fluence (*F*) for 100 nm virions within spherical particles of dried respiratory fluids with the diameters indicated (9 µm, red; 5 µm, green; 1µm, blue; 0.1 µm, black). The *S*_p_ at 260 nm are solid lines, and at 302 nm are dashed lines. Typically the *k* at 302 nm is less than the *k* at 260 nm. The particles are resting on a surface with real refractive index 1.4. The illuminating UV light propagates toward the surface in a direction perpendicular to the surface.

**Figure 2.**
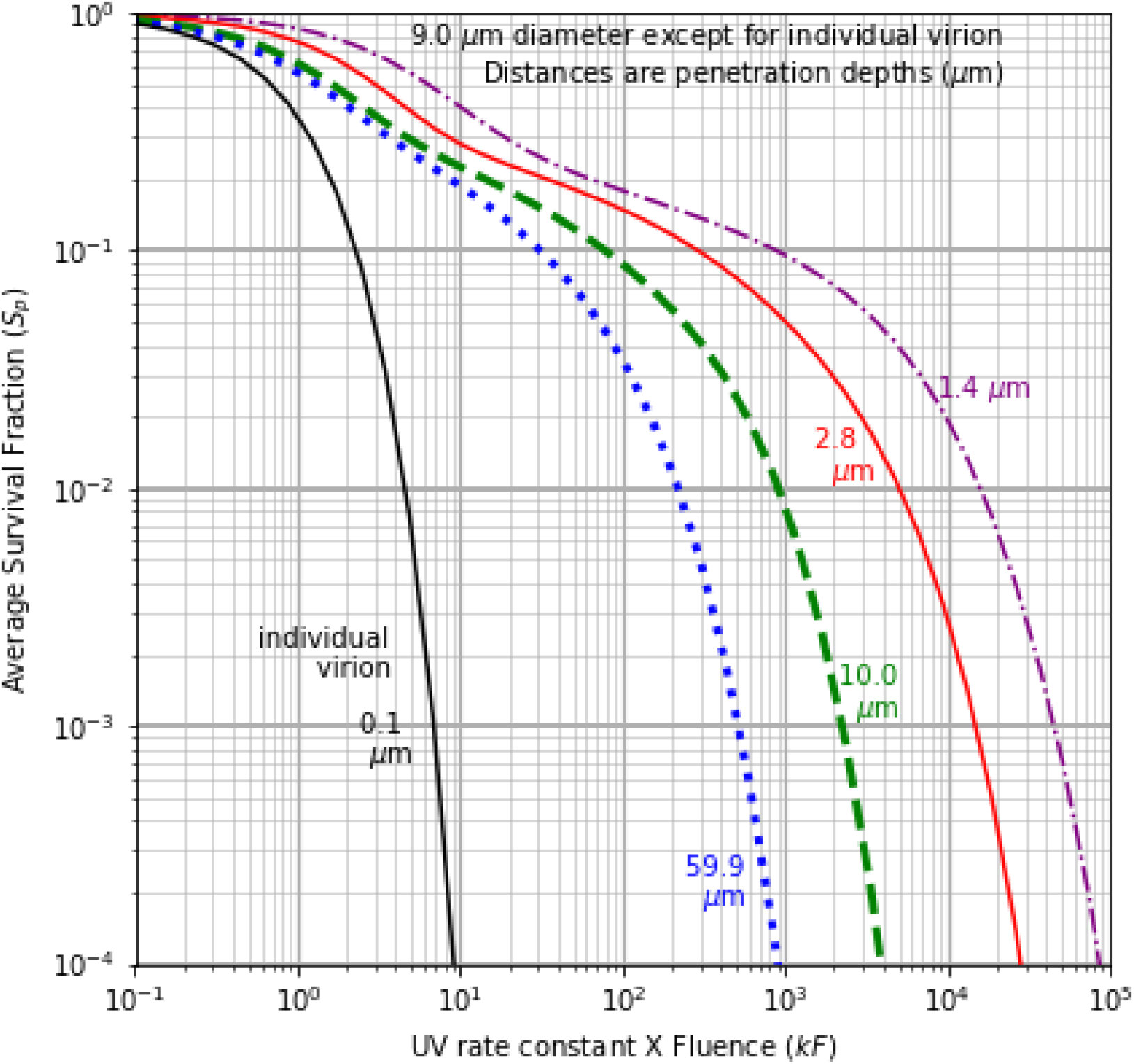
*S*_p_ vs *kF* for 9-µm particles (as in Fig. 1), except that each curve has a different penetration depth and *m*_*i*_ for its the dried respiratory fluids: 1.4 µm (*m*_*i*_ =0.01456) at 260 nm (purple); 2.8 µm (*m*_*i*_ = 0.00728) at 260 nm (red); 10 µm (*m*_*i*_ =0.00241) for a typical estimate at 302 nm (green); and, 59.9 µm (m_i_ =0.00040) for a more weakly absorbing particle at 302 nm (blue). The particle size is 9 µm except for the 0.1 µm individual virion shown for comparison.

### Average Survival Fractions vs kF are Larger in Particles with Higher Absorptivity

For a given *kF* in Fig. 1, the *S*_p_ are smaller when λ=302 nm than when λ=260 nm, because at 302 nm the particles are less absorbing of UVGI and the δ are larger. For example, when *kF*=10, the *S*_*p*_ for 1-µm particles are 0.0156 at 260 nm, but 0.00377 at 302 nm. Also when *kF*=10, the *S*_*p*_ for 5-µm particles are 0.217 at 260 nm but 0.164 at 302 nm. However, the *k* at 302 nm tend to be many times smaller than at 260 nm, and so a higher *F* (J/m^2^) is required to achieve the same *kF*.

Figure 2 illustrates how the *S*_p_ vs *kF* curves vary with the absorption of UV light by the material of the dried droplet. The curves are labeled by the *δ*. The *m*_*i*_ are in the caption and Table 1. In Fig. 2 the *m*_*i*_ were chosen to span a wider range than the estimated “typical” values used in the other figures. In Fig. 2, at 260 nm, in addition to the *δ* = 2.8 and 10 µm in Fig. 1, there is also a more strongly absorbing particle with *δ* = 1.4 µm (*m*_*i*_ = 0.01456). At 302 nm, there is also a more weakly absorbing particle with *δ* = 59.9 µm (*m*_*i*_ = 0.00040). This larger range of *δ* is used to better account for the large variations in some of the primary light-absorbing components of saliva or nasal fluids, such as, at 260 nm, nucleic acids (e.g., Poehls et al., 2018), and at 302 nm, uric acid (Hawkins et al., 1963; Riis et al., 2018, Fig. 2).

**Table 1:** Survival fraction of virions (*S*_p_) at several *kF*. Parameters include: wavelength (λ) in nm, diameter (*d*) of the large particle in µm, numbers of virions (N) in calculations, and m_i_ (imaginary component of complex refractive index). Incident angle(s) of UV illumination are specified by a zenith angle (β, in degrees), and, when needed, the azimuthal angle (α). “All” indicates averaged over all orientations. The individual virion (*d*=0.1 µm) is plotted in all figures, but only appears once in this table (top row).

| Fig | $\lambda$<br>nm | $d$<br>$\mu\text{m}$ | $N$ | $m_i$ | angle<br>degrees | $S_p$ @<br>$kF=1$ | $S_p$ @<br>$kF=10$ | $S_p$ @<br>$kF=100$ | $S_p$ @<br>$kF=1000$ | $S_p$ @ $kF=$<br>10000 |
| --- | --- | --- | --- | --- | --- | --- | --- | --- | --- | --- |
| 1 | 260 | 0.1 | 1 | 0.00728 | $\beta=0$ | 0.368 | 0.000045 | 0.000000 | 0.000000 | 0.000000 |
| 1 | 260 | 1 | 500 | 0.00728 | $\beta=0$ | 0.442 | 0.015551 | 0.000000 | 0.000000 | 0.000000 |
| 1 | 260 | 5 | 3000 | 0.00728 | $\beta=0$ | 0.671 | 0.217234 | 0.076803 | 0.004748 | 0.000000 |
| 1 | 260 | 9 | 12000 | 0.00728 | $\beta=0$ | 0.759 | 0.281645 | 0.148994 | 0.050528 | 0.002829 |
| 1 | 302 | 1 | 500 | 0.00241 | $\beta=0$ | 0.377 | 0.003765 | 0.000000 | 0.000000 | 0.000000 |
| 1 | 302 | 5 | 3000 | 0.00241 | $\beta=0$ | 0.569 | 0.163955 | 0.022992 | 0.000010 | 0.000000 |
| 1 | 302 | 9 | 12000 | 0.00241 | $\beta=0$ | 0.617 | 0.225805 | 0.087981 | 0.008403 | 0.000000 |
| 2 | 302 | 9 | 12000 | 0.0004 | $\beta=0$ | 0.565 | 0.188023 | 0.034816 | 0.000059 | 0.000000 |
| 2 | 302 | 9 | 12000 | 0.00241 | $\beta=0$ | 0.617 | 0.225805 | 0.087981 | 0.008403 | 0.000000 |
| 2 | 260 | 9 | 12000 | 0.00728 | $\beta=0$ | 0.759 | 0.281645 | 0.148994 | 0.050528 | 0.002829 |
| 2 | 260 | 9 | 12000 | 0.01456 | $\beta=0$ | 0.861 | 0.404283 | 0.179325 | 0.096010 | 0.019449 |
| 3 | 260 | 9 | 12000 | 0.00728 | $\beta=0$ | 0.759 | 0.281645 | 0.148994 | 0.050528 | 0.002829 |
| 3 | 260 | 9 | 12000 | 0.00728 | $\beta= 40$ | 0.751 | 0.227918 | 0.075299 | 0.010311 | 0.000088 |
| 3 | 260 | 9 | 12000 | 0.00728 | $\beta= 80$ | 0.718 | 0.185450 | 0.056947 | 0.012725 | 0.000289 |
| 3 | 260 | 9 | 12000 | 0.00728 | $\beta= 0,40$ | 0.754 | 0.239888 | 0.083816 | 0.011816 | 0.000162 |
| 3 | 260 | 9 | 12000 | 0.00728 | $\beta= 0,80$ | 0.734 | 0.195602 | 0.061411 | 0.013317 | 0.000342 |
| 3 | 260 | 9 | 12000 | 0.00728 | $\beta= 80,$<br>$\alpha = 0,180$ | 0.683 | 0.123693 | 0.005391 | 0.000908 | 0.000067 |
| 3 | 260 | 9 | 12000 | 0.00728 | $\beta= 80,$<br>$\alpha=0,90,180,270$ | 0.675 | 0.095058 | 0.003309 | 0.000716 | 0.000021 |
| 3 | 260 | 9 | 12000 | 0.00728 | $\beta=0 +$<br>$\beta= 80, \alpha= 0,180$ | 0.699 | 0.111898 | 0.004289 | 0.000004 | 0.000000 |
| 3 | 260 | 9 | 12000 | 0.00728 | $\beta=0 +$<br>$\beta= 80,$<br>$\alpha=0,90,180,270$ | 0.686 | 0.087636 | 0.001282 | 0.000000 | 0.000000 |
| 4 | 260 | 5 | 3000 | 0.00728 | All | 0.589 | 0.017202 | 0.000000 | 0.000000 | 0.000000 |
| 4 | 260 | 9 | 4000 | 0.00728 | All | 0.751 | 0.080112 | 0.000000 | 0.000000 | 0.000000 |

In Fig. 2, the *S*_p_ of even the least absorbing particle is many times greater than that of the individual virion. For example, even when the δ is 6.7 particle diameters (i.e., the 9-µm particle at 302 nm with δ = 59.9 µm), when *kF* = 10, the *S*_p_ for the virions in the particle (0.188) is 4200 times greater than the *S*_p_ of the individual virion (0.000045). Because the estimated absorption is so low, the increase in *S*_p_ occurs primarily because of refractive shielding. For solar wavelengths longer than the 302-nm UVGI used here, the absorption tends to be even lower, with larger δ. However, because of refractive shielding the large increase in *S*_p_ remains (as compared to that of the individual virions).

### Average Survival Fractions are Reduced with UV light from Multiple Directions

Figures 3 and 4 illustrate how illumination with beams from multiple angles can result in reductions in *S*_p_. The notation for angles follows that used in the MSTM codes (Mackowski, 2008). A line perpendicular to the surface and passing through the center of the particle and then through the planar surface defines the z-axis. The zenith angle (β) is the angle from this axis. When there is more than one illumination beam with β ≠ 0°, the azimuthal angles (α) angles between these beams are also given. In Fig. 3, when *kF* = 10, the *S*_p_ in the 9-µm particles range from 0.0876 with five widely separated UV beams, to 0.282 with one UV beam at normal incidence. For the individual virion, *S*_p_ = 0.000045. When *kF* = 100, the *S*_p_ range from 0.00128 with five widely separated UV beams to 0.149 with one UV beam with β = 0°.

**Figure 3.**
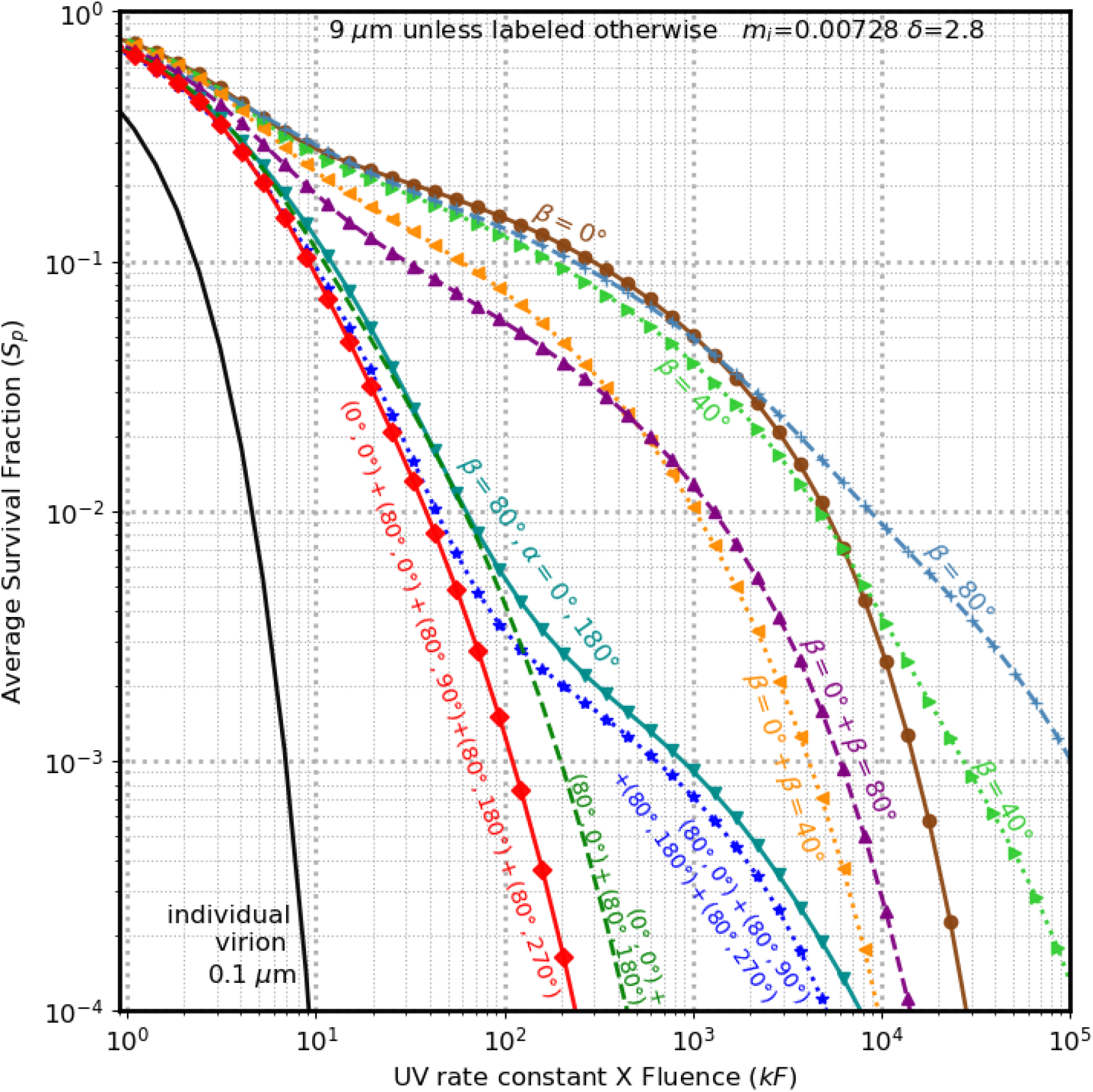
*S*_p_ vs *kF*, for 9-µm particles as in Fig. 1 (case with 260 nm illumination and δ = 2.8 µm), but with various combination of illumination beams. The three curves on the far right are each for a single illumination angle β: 0° (solid, brown circles); 40° (light green, dots and triangles; and 80° (steel blue, dashed). The next two lines have an illumination wave normal to the surface (β=0°) and a second wave at β=40° (purple), or 80° (dark yellow, dots, triangles). In each case, the total intensity illuminating the particle is the same.

**Figure 4.**
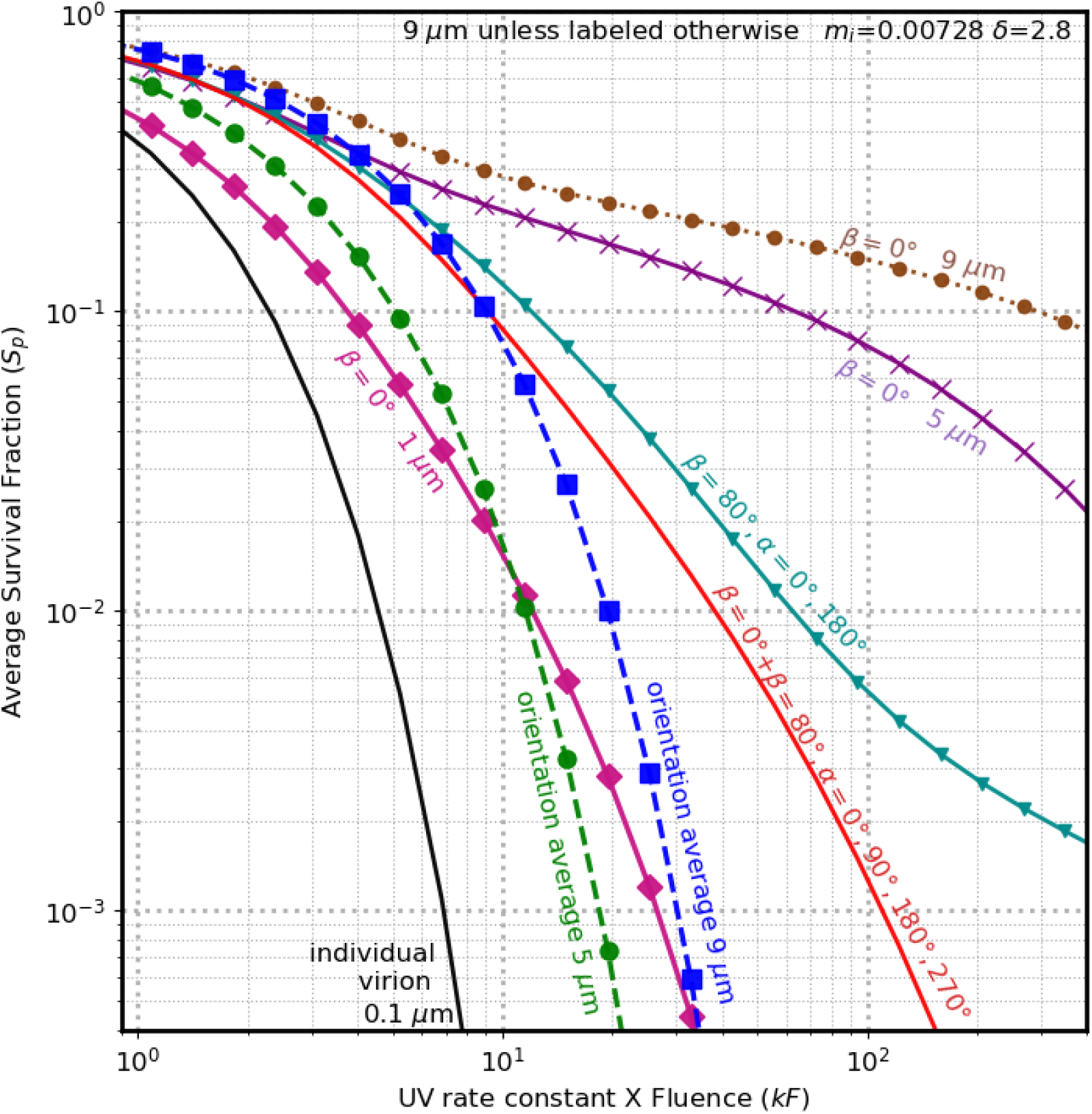
*S*_p_ vs *kF* for virions in particles as in Fig. 3 with 260-nm light, but the dashed lines are for particles illuminated with equal intensity from all directions (i.e., averaged over all orientations) with diameters (*d*) = 5 µm (green) and 9 µm (blue). Other cases are shown for particles illuminated with a wave propagating normal to the surface (β=0°): individual virions (black); *d* = 1-µm (magenta diamonds), *d* = 5-µm (purple ×), and *d* = 9-µm (brown circles with dotted line). Also shown from Fig. 3: five-beam case (solid red) and two beams with β = 80° (cyan with triangles).

In Fig. 3 the curves can be grouped into three main categories: a) Illumination with one beam only, with β = 0°, 40° or 80°. b) Illumination with multiple beams, but the differences in illumination angles are less than 90°. c) Illumination with multiple beams where at least one of the angular differences between two of the angles of incidence is greater than 90°.

With only one illumination beam the differences in *S*_p_ distributions are relatively small, but not negligible. The set of beams providing the lowest *S*_p_ includes five beams: one beam heading perpendicular to the surface (β=0°), and four beams, each with β = 80° from the vertical but approaching from different azimuthal angles, α=0°, 90°, 180°, 270°. At *kF*=100, the *S*_p_ = 0.00128 for this set. A set of four beams with β = 80° and α=0°, 90°, 180°, or 270°, (similar to the five-beam case but with no β = 0° beam), has *S*_p_ =0.00331.

Highly UV-reflective surfaces (Ryan et al., 2010; Rutala et al., 2013; Krishnamoorthy et al., 2016; Lindsley et al., 2018) can increase overall UVGI and can help in directing UVGI into shaded regions of a room. The results in Fig. 3 suggest an additional benefit of highly UV-reflective surfaces, i.e., helping to achieve illumination with multiple beams where the angular differences between the angles of incidence with respect to the surface is relatively large (see Doughty et al., (2021) for the *R*_*i*_ distributions).

In Figs. 3 and 4 the UV illumination energy is split between the beams so that the total intensity in each of the cases in Fig. 3 can be directly compared. Potential increases in the overall UV-illumination intensity that could result from generating additional beams using reflective surfaces could very likely decrease the *S*_p_. However, such calculations depend upon factors such as the positions, angles and reflectivities of the relevant surfaces, and are beyond the scope of this paper.

Figure 4 shows the orientation-averaged *S*_p_ vs *kF* for virions in 5- and 9-µm particles (dashed lines). Such orientation averaged results might be obtained with a fixed-orientation particle illuminated equally from all directions or with a particle that rotates (tumbles and spins) through all orientations with respect to one or more UV beams.

In Fig. 4 (see also the numbers in the Table) the reductions in *S*_p_ with orientation averaging are remarkable. For *kF* > 9, the all-orientations case has a lower *S*_p_ than any of the other cases with 9-µm. For 9-µm particles when *kF* =100, the *S*_p_ is less than 5 × 10^−7^ for the orientation-averaged case, but is 0.00128 for the five beam case, and 0.149 for the single beam β = 0° case. For 5-µm particles when *kF* =100, the *S*_p_ is less than 5 × 10^−7^ for orientation-averaged case, and is 0.0768 for the single beam with β = 0°.

Also in Fig. 4, when *kF* > 11 the *S*_p_ for the orientation-averaged 5-µm particles is less than the *S*_p_ for 1-µm particles (β = 0°). When *kF* > 33, the *S*_p_ for the orientation-averaged 9-µm particles is less than *S*_p_ for 1-µm particles (β = 0°). Such results may appear confusing. How can orientation averaging be so important compared to a 9-fold difference in diameter when the smaller particle is only 1-µm diameter? The reason is that in the orientation-averaged particle there are no virions that are refractively shielded well. But the wavelength is so short (260 nm), that even a 1-µm particle illuminated from one direction can have 100-nm regions that are significantly refractively shielded (e.g., Hill et al., 2015, Fig. 7a).

## DISCUSSION

### Virions in Particles Tend to have Larger Survival Fractions for a set UV Dose

Virions within UV-illuminated particles tend to have larger *S*_p_ than do individual virions, as seen in calculations for spherical particles which are homogeneous except for the virions.

Several key relations can be seen in the calculated results.

a. For particles on a surface with a given *m*_*r*_ *+ i m*_*i*_, illuminated with normally incident UVGI (zenith angle β=0) at a given wavelength, as the particle size increases the *S*_p_ increases (except for possibly at some of the lowest UV fluences) (Fig. 1). This relation occurs for both wavelengths studied (260 and 302 nm). To achieve *S*_p_ = 0.001 in 1-µm particles with 260 nm light, *kF* must be approximately 4× higher for 1-µm particles, 230× higher for the 5-µm particles and 2000X greater for the 9-µm particles, all with respect to the individual virion. At 302 nm, the increases in *kF* required are smaller: 2×, 56× and 300×, respectively, because the absorption of UV by the particle is smaller.
b. For particles on a surface illuminated with normally incident UVGI (β =0), as the penetration depth in the particles decreases the *S*_p_ increases. This relation is most clear in Fig. 2, but also seen in Fig. 1.
c. UV Illumination of particles from multiple directions, especially widely varying directions, can reduce the *S*_p_. Particles illuminated equally from all directions tend to have especially low *S*_p_ (Fig. 4). All-orientation *S*_p_ may be approximated by particles that tumble and spin for a sufficient time as they are carried within turbulent airflows through UVGI. Small particles in outdoor air on a gusty and sunny day may be examples of particles illuminated from all directions.

### Complications

#### Compositions of Coughed or Sneezed Airway Fluids are Complex and Variable

The compositions of airway fluids may vary with a person’s genetic makeup, disease state (if any), time since last drink or meal, degree of hydration, exertion level, region of the respiratory tract, and other factors. In the estimates of *m*_i_ from airway-fluid compositions used here, the molecules absorbing the most UVGI at 260-nm are DNA, RNA and nucleosides/nucleotides. In using calculated *S*_*p*_ to improve the use of UVGI in reducing the transmission of disease, a sense of the ranges of measured concentrations of UV-absorbing molecules such as nucleic acids can be beneficial. Konec̆ná et al. (2020) reported average dsDNA concentrations in whole saliva of 0.24 g/l in healthy subjects and 0.5 g/L in persons with periodontitis. However, in the supernatants from centrifuged (1600× *g*) samples they measured average concentrations of DNA of 0.054 g/l in healthy subjects and 0.07 g/l in persons with periodontitis. Fahy et al (1995) measured a range of cell-free DNA concentrations from 0.06 to 5.8 g/L (average 0.5 g/L) in patients with asthma. Pandit et al. (2013) found the extracellular RNA in the saliva (supernatant after centrifugation) ranged from 0.17 to 0.76 g/l in healthy control subjects and from 0.09 to 0.36 g/l in cancer patients. SARS-CoV-2 and other microbes can induce neutrophils in saliva to release DNA, histones and proteins to form Neutrophil Extracellular Traps (NETs) (Lachowicz-Scroggins et al., 2019; Arcanjo et al., 2020). Eosinophil Extracellular Traps (EETs) and Basophile Extracellular Traps (BETs) also contain DNA and contribute to airway defense and inflammation (Yousefi et al., 2020). Bacterial and viral DNA and RNA (Haro et al., 2020) also occur in airway fluids, but typically in lower concentrations than host DNA and RNA. To partially account for the large variability in DNA and RNA concentrations, a large range of *m*_i_ are used in the calculations here.

#### More Information is Needed on Morphologies of Dried Particles of Respiratory Fluids

In this paper the model particles are spherical and homogeneous (except for the virions), and the positions of virions within each particle are random. The MSTM can be used to model UVGI in more-complex particles than those investigated here, e.g., particles with additional spherical regions inside and outside the particle, each with its own *m*_r_ and *m*_i_. However, such modeling requires measurements or assumptions of the geometries of the particles and *m*_r_ and *m*_i_ of each of the various regions. The homogeneous-sphere-with-virions assumption is a useful case to investigate first, given the large numbers of relevant variables: *m*_r_ and *m*_i_ of the virion(s), particle and surface; size(s) of the virions and the particles; virion locations; wavelengths; number and angles of beams; presence of a surface or not. However, for more accurate modeling of the *S*_*p*_ of more complex particles, additional studies along the lines of studies by Vejerano and Marr (2017) and by Walker et al., (2021) would likely be needed.

Vejerano and Marr (2017) collected droplets of artificial saliva on a superhydrophobic surface and used optical microscopy to examine the particles as they dried as the RH was lowered. In their figures, particles that were less than roughly 20 µm appear to us as approximately spherical, but inhomogeneous. The nonsphericities appear to arise from the crystallization/precipitation of different materials in different locations. Walker et al., (2021, their figure 5c) collected dried particles of artificial saliva on a surface and examined them with scanning electron microscopy. The particles generally look smooth on the order of the apparent resolution, except for some unclear features. Walker et al. (2021) also state that light scattering patterns indicate that the particles are inhomogeneous. These are the only studies (to the authors’ knowledge) with as much information related to morphologies of small particles from dried saliva or respiratory fluids with sizes in the range of interest (e.g., Fennelly, 2020). However, there are uncertainties in how to apply the observations of Vejerano and Marr, (2017) and Walker et al. (2021) to modeling of *S*_*p*_ of dried saliva or respiratory particles in the 0.5- to 9-µm size range.

One reason for the uncertainty is that the artificial saliva particles studied by Vejerano and Marr, (2017) and Walker et al. (2021) are mostly > 20 µm. In contrast, the particles emphasized here are 0.1- to 9-µm; larger particles are significant on a per-mass basis (Jarvis, 2020), but tend to deposit relatively quickly from air. As droplets of respiratory fluid dry, some solutes may crystalize into different regions, resulting in inhomogeneous and possibly nonspherical particles. As water evaporates from the surface, concentration gradients form within the drying droplets and the droplets become more viscous. The distributions and gradients of solutes depend on the diffusion coefficients of the solutes, and the temperature- and humidity-dependent rates of evaporation. A given concentration difference requires a higher concentration gradient in a 1-µm particle than in a 20-µm particle.

Another reason for uncertainty in applying the measured results is that respiratory fluids tend to be more complex than the artificial fluids used in the studies mentioned. Differences between human and model saliva, and between gastric and submaxillary mucin, are not insignificant (Sarkar et al., 2019). Because of the additional complexity of actual saliva and other respiratory fluids (containing DNA, RNA, multiple mucins and other proteins, carbohydrates, etc.), there is cause to wonder if crystallization/precipitation may be more likely to occur as smaller crystals in dried coughed/sneezed droplets, as compared with fewer larger crystals in the artificial salivas. Vejerano and Marr (2017) used an aqueous solution of gastric mucin, NaCl, and a phospholipid surfactant. Walker et al. (2020), following (Woo et al., 2012), used an aqueous solution of Na^+^, K^+^, Mg^2+^, Ca^2+^, Cl^-^, HCO_3_ ^-^, H_2_PO_4_ ^-^, SCN^-^, amylase, mucin from porcine stomach, and a 1-to-1000 dilution of Dulbecco’s modified minimal essential medium, which adds very small concentrations of glucose, amino acids, vitamins, cofactors, and sulfate. Neither includes nucleic acids. Large molecules such as DNA and RNA, not included in the artificial saliva, may affect the diffusion rates of various solutes (Dix and Verkman, 2008) or particles (Linssen et al., 2021) as they respond to the concentration gradients generated by the evaporation of water from the surface of the droplet/particle.

## CONCLUSIONS

Average survival fractions (*S*_p_) of virions in particles of dried respiratory fluids exposed to UVGI can be much larger than survival fractions of virions not within such particles, as illustrated in Figs. 1-4. The *S*_p_ were calculated using UV intensities within virions calculated with the MSTM, an exact method (within numerical error). The model particles are spherical and homogeneous, except for the virions. The particles may be on a surface and may be illuminated with any number of incident beams.

The key findings are:

a. The *S*_p_ increases as the particle size increases, where the smallest relevant particle is an individual virion (Figs. 1 and 4).
b. The *S*_p_ increases as the UV-absorptivity of the material comprising the particle increases (Figs. 1 and 2). However, even when the material of the particle absorbs very little UV light, the *S*_p_ increases as the particle size increases.
c. The *S*_p_ tends to decrease as more UV beams, from more widely separated directions, illuminate the particle, where the total UV energy is the same for each case (Figs. 3 and 4). In the limiting case of equal illumination from all angles, the *S*_p_ tend to be especially low (Fig. 4). This case is equivalent to the one where particles rotate (spin and tumble) through all orientations with respect to the illumination beams during their time of exposure to UVGI.
d. Increases in *S*_p_ for virions in particles can be mitigated by using more intense UVGI sources, and/or more UVGI sources, and/or with highly UV-reflective surfaces (Ryan et al., 2010; Rutala et al., 2013; Lindsley et al., 2018). An advantage of using highly UV-reflective surfaces is that in addition to increasing the UV intensity, they can also be positioned to increase the numbers of directions from which particles are illuminated, and the angular separation between the various angles of illumination.

Detailed, accurate modeling of the *S*_p_ of virions in particles requires knowledge of the morphologies of the virion-containing particles, i.e., the 3-D shapes, positions, and compositions or optical properties of each region of the particle.

## RECOMMENDATIONS

In designing, testing and using UVGI systems to inactivate viruses, recognize that virions within particles larger than a very few UV wavelengths tend to have larger survival fractions than do individual virions, and that the survival fractions increase with particle size.

In evaluating UVGI systems for efficacy of inactivation, test for inactivation with virion-containing particles having sizes and compositions representative of the particles that need to be inactivated. For example, if tests were done only with virion-containing particles smaller than 1 µm, but the system may be used with a significant fraction of particles as large as 4 to 5 µm, then recognize that the results shown here suggest that the measured survival fractions will very likely be higher for the 4- to 5-µm particles.

In designing and deploying UVGI systems for virions in particles sufficiently large- and/or absorptive to shield virions, consider, to the extent possible, systems that illuminate particles from many, widely separated, angles (see Figs. 3 and 4). Consider employing systems with surfaces that are highly reflective at the relevant UV wavelengths, because such surfaces can increase both the UV intensities and the ranges of angles of incidence of the UV illumination, and thereby decrease the survival fractions. For systems in which particles of all the relevant sizes spin and tumble through all orientations as they are being illuminated, the need for illumination from many directions, including reflected beams, is not necessary.

## Data Availability

The codes to make the figures and calculate the numbers in the table are available upon request from.
Codes to calculate numbers that were used in calculating the values and figures shown here are available as described in the paper by David Doughty, Steven Hill and Dan Mackowski, "Viruses such as SARS-CoV-2 can be partially shielded from UV radiation when in particles 1 generated by sneezing or coughing: Numerical simulations", J Quant Spectrosc Radiative Transfer 2021. https://doi.org/10.1016/j.jqsrt.2020.107489

